# A highly specific assay for the detection of SARS-CoV-2-reactive CD4+ and CD8+ T cells in COVID-19 patients

**DOI:** 10.1101/2020.08.10.20150060

**Authors:** Henning Zelba, David Worbs, Johannes Harter, Natalia Pieper, Christina Kyzirakos-Feger, Simone Kayser, Marcel Seibold, Oliver Bartsch, Jiri Ködding, Saskia Biskup

**Author notes:** **<u>Corresponding author:</u>** Henning Zelba, Fon.

## Abstract

Gaining detailed insights into the role of host immune responses in viral clearance is critical for understanding COVID-19 pathogenesis and future treatment strategies. While studies analyzing humoral immune responses against SARS-CoV-2 were available rather early during the pandemic, cellular immunity came into focus of investigations just recently.

For the present work, we have adapted a protocol, designed for the detection of rare neoantigen-specific Memory T cells in cancer patients for studying cellular immune responses against SARS-CoV-2. Both, CD4+ and CD8+ T cells were detected after 6 days of in vitro expansion using overlapping peptide libraries representing the whole viral protein. The assay readout was an Intracellular cytokine staining and flow cytometric analysis detecting four functional markers simultaneously (CD154, TNF, IL-2, IFN-γ).

We were able to detect SARS-CoV-2-specific T cells in 9 of 9 COVID-19 patients with mild symptoms. All patients had reactive T cells against at least one of 12 analyzed viral antigens and all patients had Spike-specific T cells. While some antigens were detected by CD4+ and CD8+ T cells, Membrane protein was mainly recognized by CD4+ T cells. Strikingly, we were not able to detect SARS-CoV-2-specific T cells in 9 unexposed healthy individuals.

We are presenting a highly specific protocol for the detection of SARS-CoV-2-reactive T cells. Our data confirmed the important role of cellular immune responses in understanding SARS-CoV-2 clearance. We showed that Spike is the most immunogenic antigen. We have introduced Membrane protein as interesting target for studying humoral immune responses in convalescent COVID-19 patients.

## Introduction

The novel Severe Acute Respiratory Syndrome Coronavirus 2 (SARS-CoV-2), the causative virus of a respiratory disease termed COVID-19, is a betacoronavirus related to Severe Acute Respiratory Syndrome Coronavirus (SARS-CoV) and Middle East Respiratory Syndrome Coronavirus (MERS-CoV) (1-6).

The appearance of SARS-CoV-2 has led to a rapidly spreading pandemic. First cases occurred in December 2019 and until the 1^st^ of July, more than 500,000 deaths and 10,000,000 cases of SARS-CoV-2 infection have been reported worldwide (Dong et al., 2020, Wu and McGoogan, 2020; Johns Hopkins University).

Several attributes of SARS-CoV-2 have contributed to its rapid spread. These characteristics include the capability to transmit already during the asymptomatic phase of infection^11,12^ and its variable incubation time of about 3–14 days. Furthermore, even asymptomatic and presymptomatic SARS-CoV-2-infected individuals can produce high viral loads sufficient for human-to-human transmission (7-9).

Diagnosis of COVID-19 is routinely achieved by detection of SARS-CoV-2 RNA in nasopharyngeal swabs via RT-qPCR (10), however, even symptomatic SARS-CoV-2 infections frequently remain unrecognized. When symptomatic, COVID-19 can range from a mild common flu-like sickness in about 85% to a severe respiratory disease in about 15% of affected patients (11, 12). Mild COVID-19 is characterized by ageusia, fever, sore throat, cough and mild pneumonia. Severe disease features strong dyspnea, hypoxia and radiographic evidence of lung involvement. Ultimately, severe COVID-19 can lead to acute respiratory distress syndrome (ARDS) with respiratory failure and multiorgan dysfunction (13).

Similar to other coronaviruses, SARS-CoV-2 infection leads to an activation of the innate and adaptive immune system. Protective immunity is meant to be achieved on the one hand by activated B-cells, comprising transient immunoglobulin M (IgM) and immunoglobulin A (IgA), and persisting immunoglobulin G (IgG) responses against the virus (Guo et al. posted on medRxiv). On the other hand, activated CD4+ and CD8+ T cells contribute to both the clearance of the acute infection, and protective immunity against reinfection by establishing immunological memory (14). Various studies showed that a high percentage of recovered COVID-19 patients including asymptomatic patients have IgA/IgM and/or IgG antibodies against SARS-CoV-2 and that their convalescent plasma has neutralizing capability (15, 16)(Wajnberg et al. posted on medRxiv, Fafi-Kremer et al. posted on medRxiv). Furthermore, different research groups used convalescent plasma from recovered COVID-19 patients for the treatment of patients with severe illness (17) (Liu et al. posted on medRxiv).

Regarding T cell immunity, several reports showed that SARS-CoV-2-specific CD4+ and CD8+ T cells are present in COVID-19 patients (18, 19)(Braun et al. posted on medRxiv, Oja et al. bioRxiv, Gallais et al. posted on medRxiv). Most of these trials were using overlapping peptides for studying T cells responses to work HLA-independently. Interestingly, almost all studies were able to detect SARS-CoV-2-specific T cells also in unexposed and/or PCR negative patients as well as historical controls that were not able to be in contact with the novel coronavirus SARS-CoV-2. This finding is explained by the possibility that pre-existing S-reactive T cells in seronegative individuals represent cross-reactive clones against the Spike-protein, that might have been acquired as a result of previous exposure to other seasonal human Coronaviruses (18) (Braun et al. posted on medRxiv).

In the present study, we applied a protocol which was initially designed for the detection of rare tumor-associated antigen (TAA)-specific or neoantigen-specific T cells in late-stage cancer patients (20-22). We expanded cryopreserved PBMCs with overlapping peptide libraries and detect viral-specific T cells by an Intracellular Cytokine Staining which allowed us to functionally characterize SARS-CoV-2-specific CD4+ and CD8+ Memory T cells.

## Material and Methods

### Study subjects

Nine unexposed donors and nine COVID-19 patients were included in the study. COVID-19 patients included individuals that were tested positive for SARS-CoV-2 RNA in nasopharyngeal swabs by RT-qPCR and symptomatic relatives of those individuals. All patients showed Spike (S1 domain) specific IgG antibodies using ELISA. Unexposed donors included individuals tested negative for SARS-CoV-2 by RT-qPCR and asymptomatic individuals with no contact to SARS-CoV-2 infected persons. None of the unexposed individuals showed S1-specific antibodies. Blood was collected between February and June 2020 in Tübingen, Germany. All participants provided written informed consent. The study was approved by the “Ethik-Kommission der Landesärztekammer Baden-Württemberg” (F- 2020-111).

### Peripheral blood mononuclear cells isolation

Whole blood was drawn in Sodium-Heparin collection tubes (Sarstedt). Peripheral blood mononuclear cells (PBMCs) were isolated by density gradient centrifugation using Biocoll Separation Solution (Biochrom). After isolation, PBMCs were washed and cryopreserved using freezing medium containing 10% DMSO (VWR) until usage.

### Overlapping peptide libraries

Protein-spanning overlapping peptides were obtained for 12 SARS-CoV-2 antigens (PepMiX; JPT). Antigens included Spike (delivered in two subpools of 158 & 157 peptides; abbreviation: S1 and S2), Nucleocapsid (NCAP; 102 peptides), Protein 3a (AP3A; 66 peptides), Envelope small membrane protein (VEMP; 16 peptides), Membrane protein (VME1; 53 peptides), Uncharacterized protein 14 (Y14; 16 peptides), ORF10 Protein (ORF10; 7 peptides), ORF9b Protein (ORF9b; 22 peptides), Non-Structural protein 6 (NS6; 13 peptides), Non-Structural protein 7A (NS7A; 28 peptides), Non-Structural protein 7B (NS7B; 8 peptides), Non-Structural protein 8 (NS8; 28 peptides).

### Detection of SARS-CoV-2-specific T cells

After thawing, PBMCs were dissolved in TexMACS medium (Miltenyi) containing 3 μg/ml DNAse I (Sigma-Aldrich) and were cultivated in a standard incubator (Eppendorf; 37°C; 5% CO2) for 12 hours. After this initial pre-incubation phase, cells were washed and re-sowed in TEXMACS medium containing 1% Penicillin-Streptomycin (Sigma-Aldrich) in a 48 well-plate. Overlapping peptides (PepMix) were added at a concentration of 1 μg/ml each. Cells were cultivated together with peptides for 6 days. After the first 24 hours of cultivation with peptides, 10 U/ml IL-2 (Miltenyi Biotec) and 10 ng/ml IL-7 (Miltenyi Biotec) were added. Medium was changed when necessary. After 5 days of cultivation with peptides, medium without cytokines was added. After 6 days of cultivation with peptides, expanded cells of each well were collected, washed and re-sowed in two wells of a 96 well-plate. The first well was restimulated with corresponding PepMix whereas the second well remained unstimulated. Golgi inhibitors (Golgi-Plug; BD biosciences) were added at a concentration of 1 μl/ml and cells were cultivated for 12 ± 2 hours.

### Short-term stimulation test

After thawing and pre-incubation, PBMCs were washed and re-sowed in TEXMACS medium containing 1% Penicillin-Streptomycin (Sigma-Aldrich) in a 96 well-plate. Overlapping peptides representing the whole antigen (PepMix; JPT) were added at a concentration of 1 μg/ml each. Cells were cultivated together with peptides for 12 ± 2 hours in presence of Golgi-Plug (BD biosciences) at a concentration of 1 μl/ml.

### Flow cytometry

For both approaches, the final readout was an Intracellular Cytokine Staining (ICS). After cultivation, cells were washed twice followed by extracellular staining with fluorochrome-conjugated antibodies titrated to their optimal concentrations: CD3-BV785 (clone UCHT1; BioLegend), CD4-FITC (clone RPA-T4; BioLegend), CD8-APC/Cyanine (clone SK1; BioLegend), Zombi Aqua Dye (BioLegend).

After extracellular staining, cells were fixed and permeabilized (BD biosciences), followed by an intracellular staining with the following antibodies: IFN-BV421 (clone 4S.B3; BioLegend), TNF-AlexaFluor700 (clone MAb11; BioLegend), IL-2-PE/Cy7 (clone MQ1- 17H12; BioLegend) and CD154 - BV711 (clone 24-31; BioLegend). Finally, cells were measured on a Novocyte 3005R cytometer (ACEA biosciences).

### Statistics

Data were analyzed using FlowJo version 10.5.3 (FlowJo LLC). After removal of duplicates using the forward-scatter area versus forward-scatter height plot, dead cells were excluded by gating on the Zombie Aqua-negative cells. Next, CD4+ and CD8+ cells were gated within viable CD3+ lymphocytes and analyzed separately for each functional marker (CD154, IFN-γ, TNF, IL-2). A detailed gating strategy can be found in Supplementary Table I. For each functional marker, we evaluated the percentage of positive cells among all gated CD4+ and CD8+ T cells in sample one (restimulated) and sample two (not restimulated). The donor was defined as having antigen-specific T cells if the stimulation index was ≥ 2 (sample one divided by sample two) for any functional marker in either CD8+ or CD4+ T cells. The frequency of cytokine-producing cells (or CD154 expressing) cells was determined by subtracting the frequency of cytokine-producing cells in sample two (not restimulated) from sample one (restimulated). All experiments were performed centrally by one investigator (DW) and analyzed by another investigator (HZ).

Frequencies of IFN-γ+ T cells within COVID-19 patients and unexposed individuals was compared with unpaired Mann-Whitney test using Prism 8.4.2 (Graphpad Software).

## Results

### Demographic and clinical characteristics of the patient cohort

Nine unexposed donors (4 female) and nine COVID-19 patients (5 female) were included in the study. Detailed characteristics are indicated in Table I. All COVID-19 patients had mild, flu-like symptoms and none of them was hospitalized. The most common symptom was limb aches followed by sore throat and fever. The mean time between symptoms/PCR+ and end of symptoms/PCR- was 16 ± 8 days. The mean time between end of symptoms/PCR- and blood draw was 34 ± 27 days.

**Table I.**
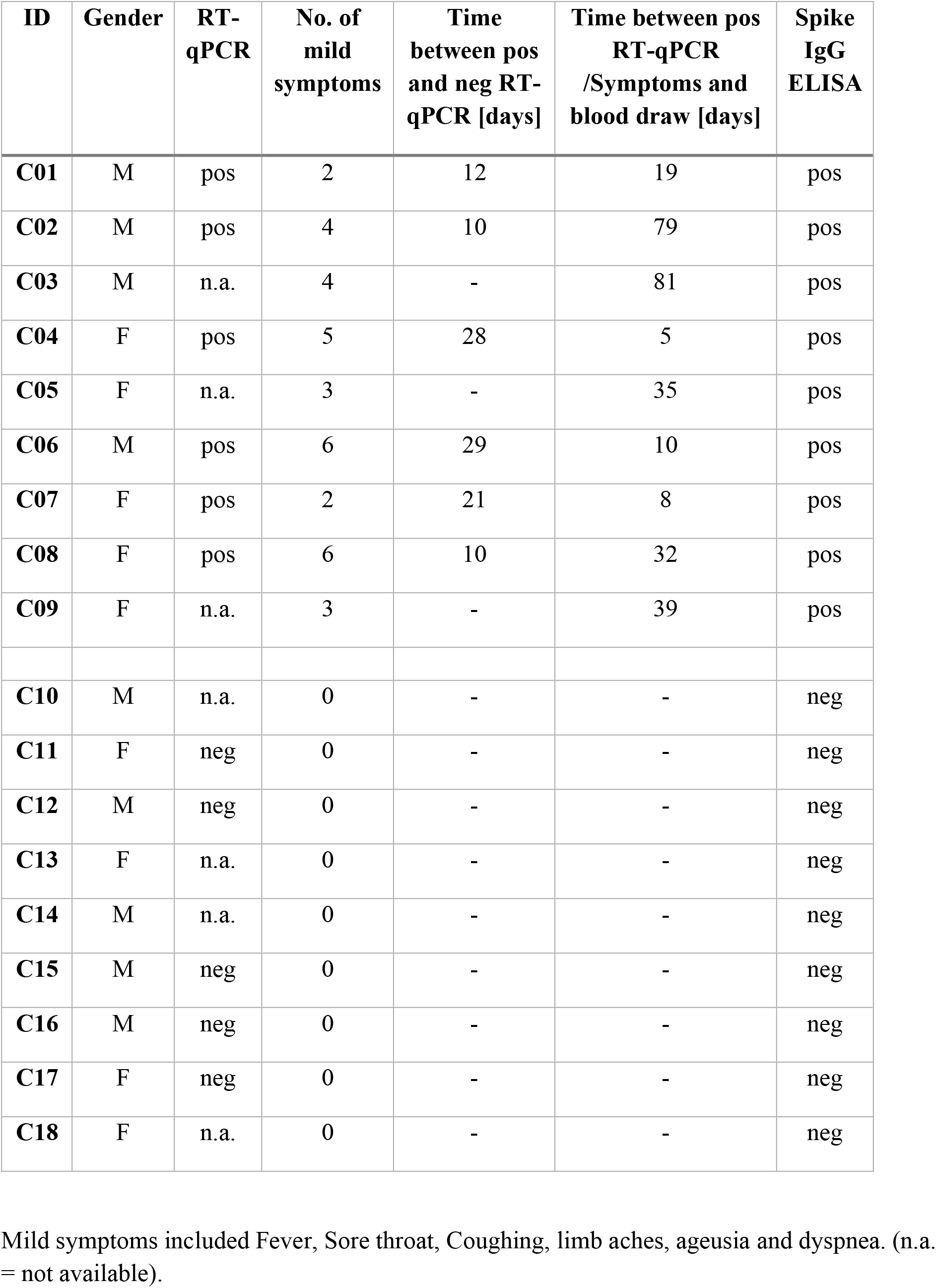
Patient characteristics

| **ID** | **Gender** | **RT-qPCR** | **No. of mild symptoms** | **Time between pos and neg RT-qPCR [days]** | **Time between pos RT-qPCR /Symptoms and blood draw [days]** | **Spike IgG ELISA** |
| --- | --- | --- | --- | --- | --- | --- |
| **C01** | M | pos | 2 | 12 | 19 | pos |
| **C02** | M | pos | 4 | 10 | 79 | pos |
| **C03** | M | n.a. | 4 | - | 81 | pos |
| **C04** | F | pos | 5 | 28 | 5 | pos |
| **C05** | F | n.a. | 3 | - | 35 | pos |
| **C06** | M | pos | 6 | 29 | 10 | pos |
| **C07** | F | pos | 2 | 21 | 8 | pos |
| **C08** | F | pos | 6 | 10 | 32 | pos |
| **C09** | F | n.a. | 3 | - | 39 | pos |
| **C10** | M | n.a. | 0 | - | - | neg |
| **C11** | F | neg | 0 | - | - | neg |
| **C12** | M | neg | 0 | - | - | neg |
| **C13** | F | n.a. | 0 | - | - | neg |
| **C14** | M | n.a. | 0 | - | - | neg |
| **C15** | M | neg | 0 | - | - | neg |
| **C16** | M | neg | 0 | - | - | neg |
| **C17** | F | neg | 0 | - | - | neg |
| **C18** | F | n.a. | 0 | - | - | neg |

Mild symptoms included Fever, Sore throat, Coughing, limb aches, ageusia and dyspnea. (n.a. = not available).

### Presence of SARS-CoV-2-specific T cells

When we analyzed flow cytometry data regardless of phenotype (CD4+ vs. CD8+ T cells) and function (CD154, IFN-γ, TNF, IL-2) according to predefined criteria (21), all COVID-19 patients had specific T cells against at least one viral antigen. Eight of 9 patients had specific T cells against more than one SARS-CoV-2 antigen. In contrast, none of the unexposed donors had specific T cells against any of the tested antigens. The most common SARS- COV-2 antigen recognized by the patients’ T cells was Spike, where Spike1-specific T cells were detected in 9 of 9 patients and Spike2-specific T cells were detected in 7 of 9 patients. Seven of 9 patients had T cells able to recognize the Nucleocapsid- and Membrane-protein. In two of 9 patients, we detected ORF10- and Protein 3a-specific T cells. One patient had ORF9b-specific T cells. None of the COVID-19 patients had T cells against any other viral antigen (Figure 1A).

**Figure 1:**
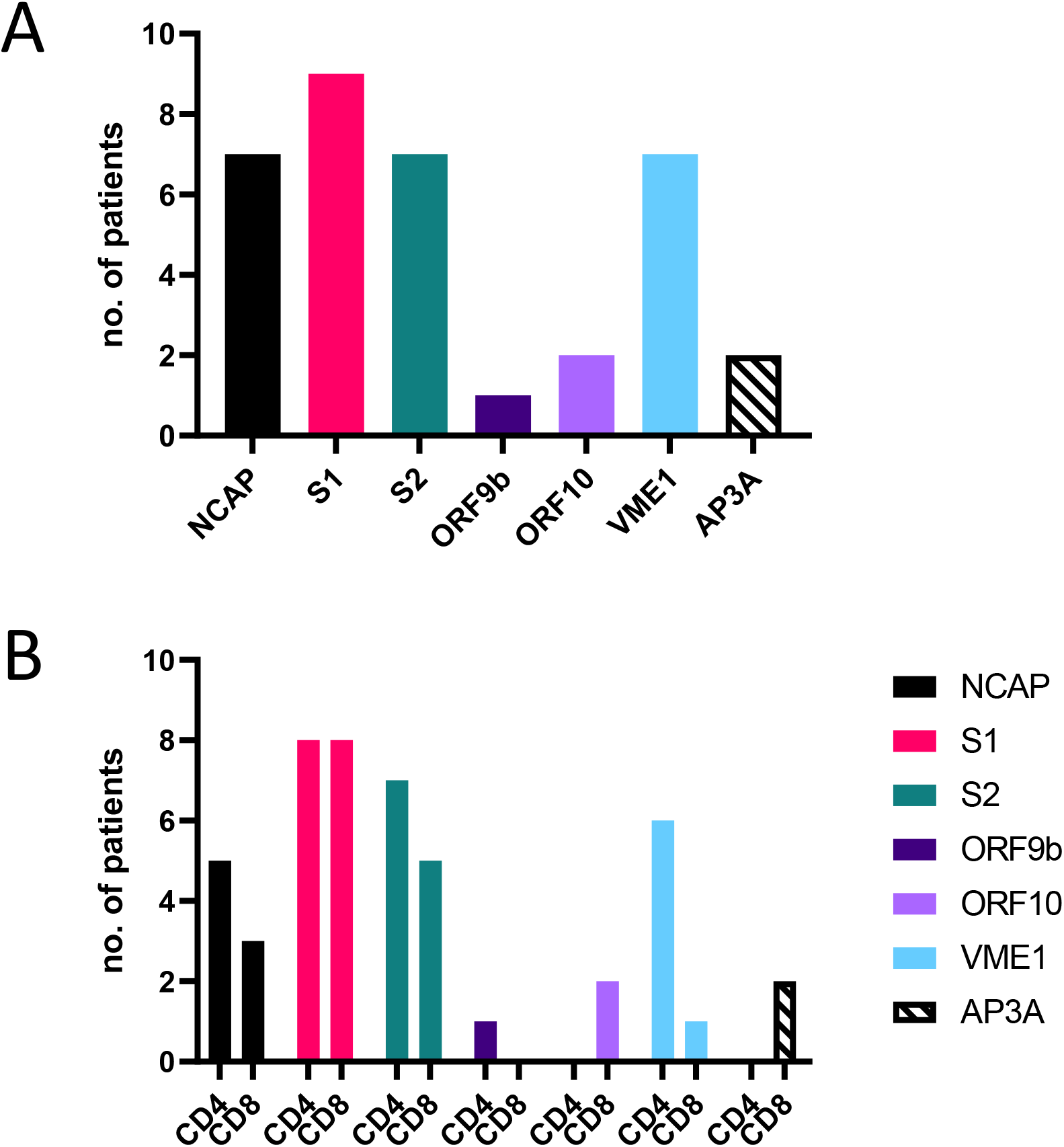
SARS-CoV-2-specific T cells in recovered COVID-19 patients (n=9) Number of patients with SARS-CoV-2-specific T cells according to our predefined response criteria regardless of phenotype (A) or separately for CD4+ and CD8+ T cells (B) after 6 days of in vitro expansion. SARS-CoV-2-reactive T cells were not detected in unexposed individuals.

### Phenotype of SARS-CoV-2-specific T cells

The Spike1 antigen was equally recognized by CD4+ and CD8+ T cells (both 8 of 9). This was similar for the Spike2 antigen (7 of 9 patients had CD4+ T cells; 5 of 9 had CD8+ T cells). A majority of Nucleocapsid-specific T cells was CD4+ (5 of 9 patients versus 3 of 9 CD8+ T cells). Interestingly, the Membrane antigen was mainly detected by CD4+ T cells (6 of 9 as compared to 1 of 9 within CD8+ T cells) whereas Protein 3a-specific T cells were exclusively CD8+ (2 of 9 patients). The immunogenicity of the Membrane protein is comparable to the Nucleocapsid protein, which introduces the Membrane antigen as interesting target for future antibody studies (Figure 1B).

### Frequency of SARS-CoV-2-specific T cells

Furthermore, we compared the frequency of IFN-γ producing SARS-CoV-2 specific CD4+ and CD8+ T cells within COVID-19 patients and unexposed individuals. Looking at CD4+ IFN-γ+ T cells, highest median frequencies of specific T cells within COVID-19 patients were observed for S2 antigen (0.43% IQR: 0.19% - 0.97%) followed by S1 (0.37%; IQR: 0.13% - 0.93%) and NCAP (0.2%; IQR: 0.02% - 1.1%). Within unexposed individuals, these frequencies were significantly reduced (S2: 0.08%; IQR: -0.06% - 0.13%. S1: -0.08%; IQR: - 0.19% - 0.04%. NCAP: -0.01; IQR: -0.1% - 0.02%).

Within CD8+ T cells, highest median frequencies within COVID-19 patients were observed for the S1 antigen (0.50% IQR: 0.21% - 2.73%) followed by S2 (0.18%; IQR: 0.15% - 0.84%). Again, frequencies within unexposed individuals were significantly decreased (S1: - 0.01%; IQR: -0.07% - 0.04%. S2: -0.03%; IQR: -0.17% - 0.09%. NCAP: 0.01; IQR: -0.08% - 0.22%) (Figure 2).

**Figure 2:**
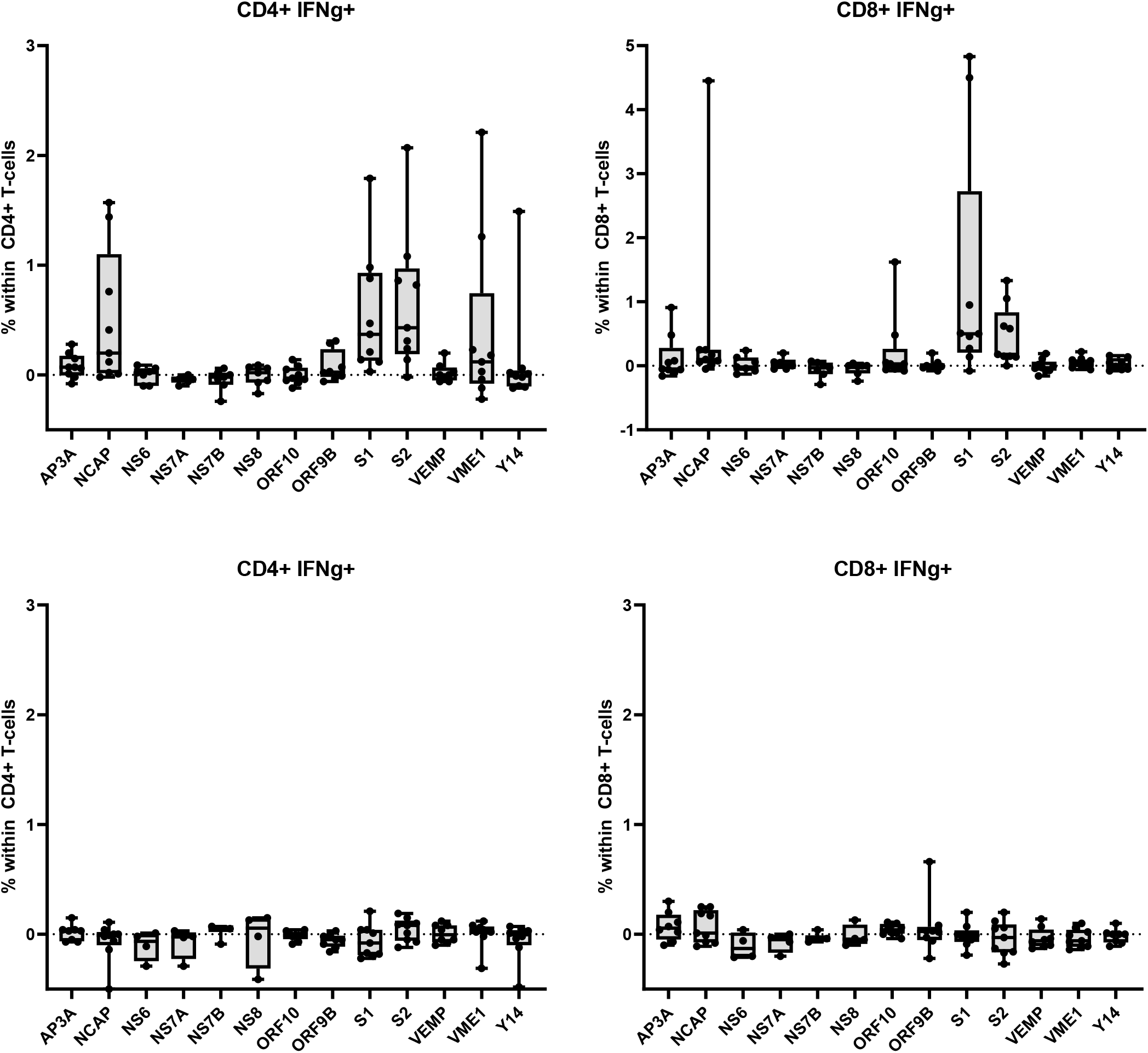
Differences between exposed (n=9) and unexposed donors (n=9) after long-term stimulation. Frequency of IFN-γ+ CD4+ (left) and CD8+ T cells (right) responding upon viral antigen stimulation within recovered COVID-19 patients (upper row) and unexposed individuals (lower row). The frequency was calculated by subtracting the frequency of IFN-γ-producing cells in sample two (not restimulated cells) from sample one (restimulated cells) after 6 days of in vitro expansion. Box-Whiskers plot (Line = Median; Box = 25th to 75th percentiles; Whiskers Min to Max; all individual shown as dots).

### Differences between long- and short-term stimulation

In contrast to our results, recent publications reported medium to high frequency of unexposed individuals with SARS-CoV-2-specific T cells. To investigate if the observed differences were donor-dependent, we performed the same set-up within the same donors using a short-term stimulation. When we analyzed flow cytometry data regardless of phenotype and function, we detected SARS-CoV-2-specific T cells in 9 of 9 COVID-19 patients. However, we have also detected SARS-CoV-2-specific T cells in 8 of 9 unexposed individuals (Table II).

**Table II.**
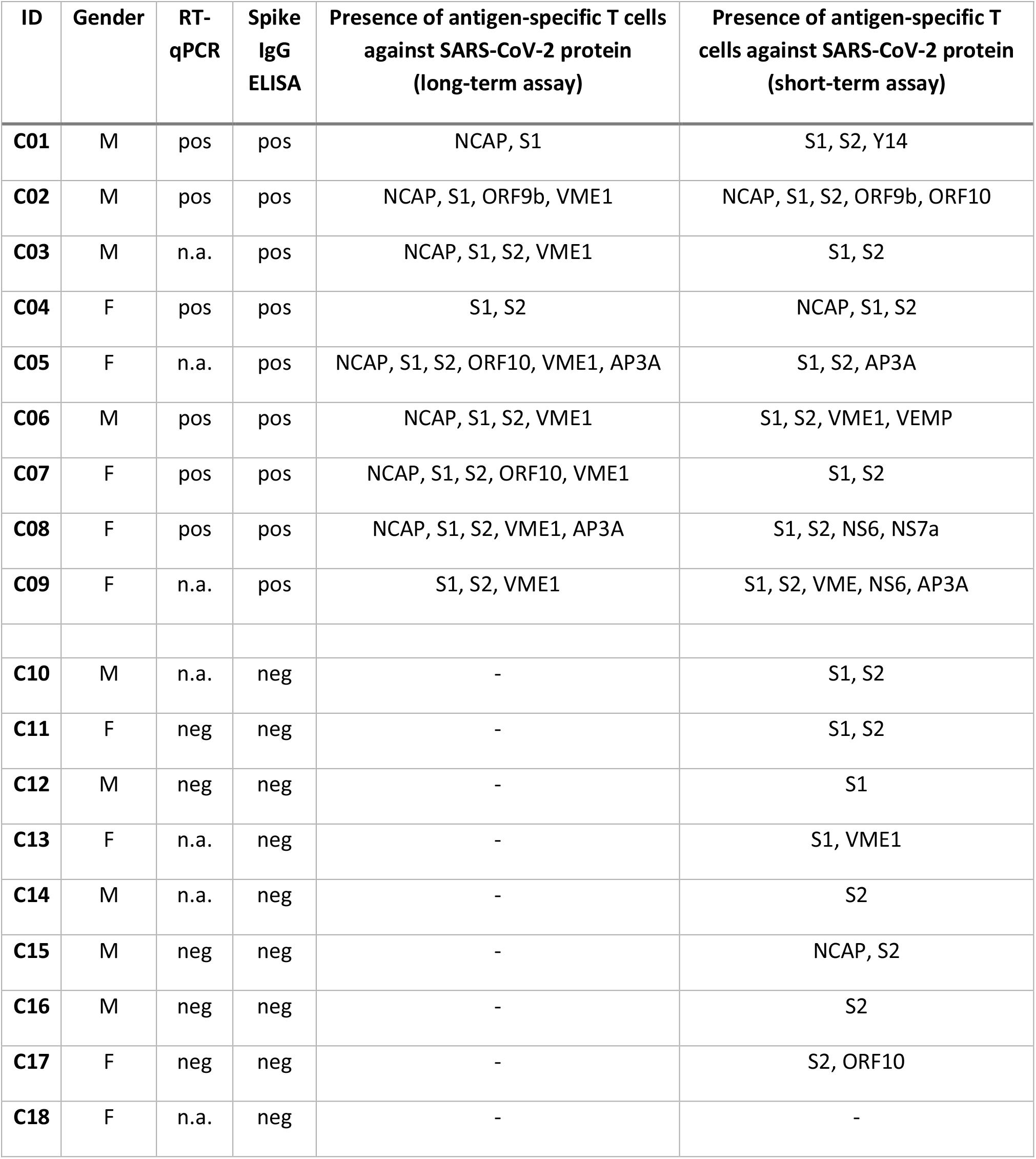
Overview of SARS-CoV-2-specific T cells using our standard method and the commonly used short-term stimulation

Regarding frequencies of specific T cells, highest median CD4+ IFN-γ+ T cells within COVID-19 patients were observed for S1 antigen (3.07% IQR: 0.18% - 4.83%) followed by S2 antigen (1.92%; IQR: 0.89% - 8.75%). Frequencies were similar for unexposed individuals in this setting (S1: 2.83%; IQR: 0.35% - 5.13%. S2: 2.44%; IQR: 1.27% - 4.37%) (Figure 3).

**Figure 3:**
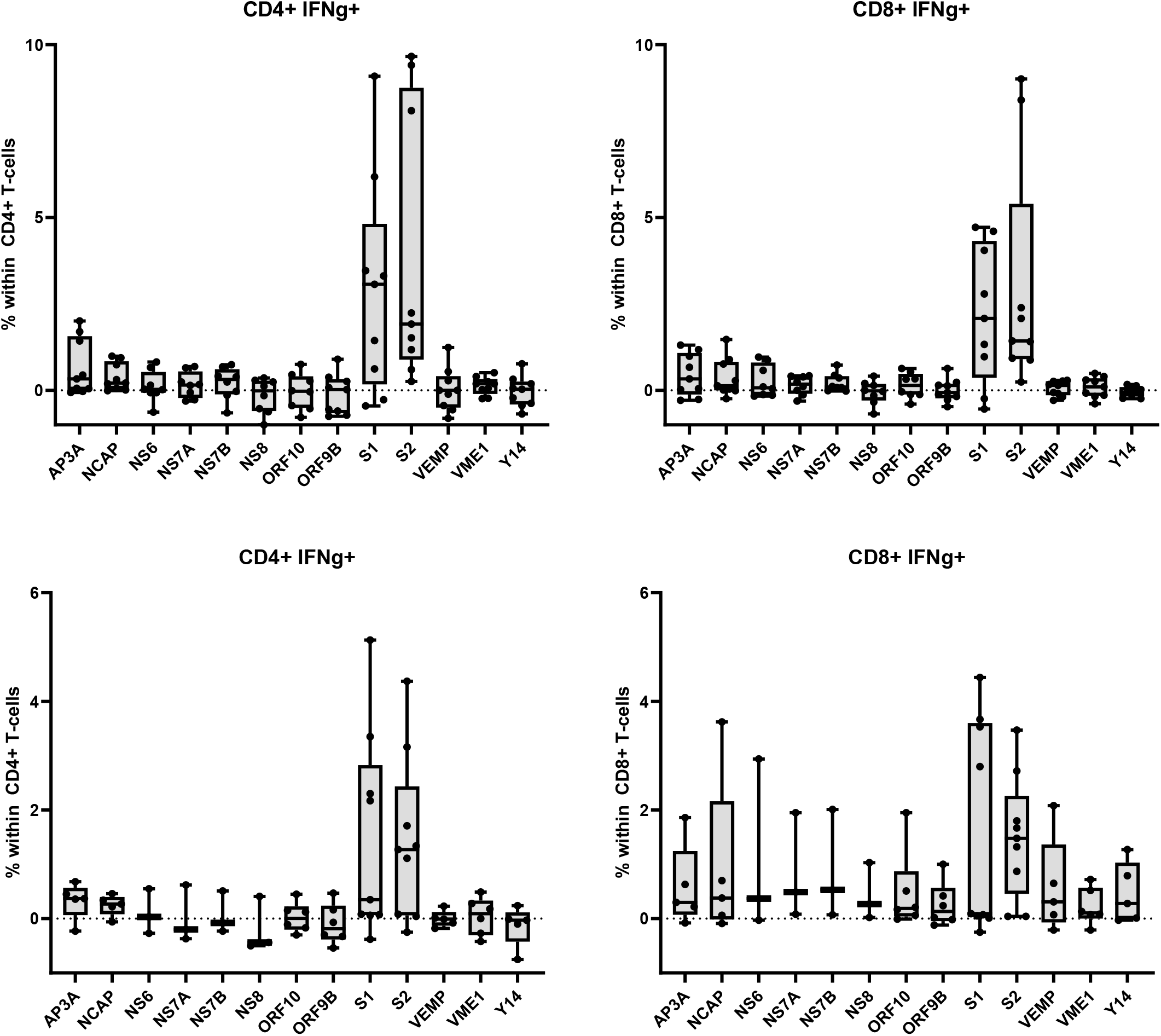
Differences between exposed (n=9) and unexposed donors (n=9) after short-term stimulation. Results from the short-term stimulation. Frequency of IFN-γ+ CD4+ (left) and CD8+ T cells (right) responding upon viral antigen stimulation within recovered COVID-19 patients (upper row) and unexposed individuals (lower row). The frequency was calculated by subtracting the frequency of IFN-γ-producing cells in sample two (not restimulated cells) from sample one (restimulated cells) after 12 hours of stimulation. Box-Whiskers plot (Line = Median; Box = 25th to 75th percentiles; Whiskers according to Tukey and outliers shown as individual dots).

Looking at CD8+ T cells, highest median frequencies within COVID-19 patients were observed for the S1 antigen (2.08% IQR: 0.37% - 4.33%) followed by S2 (1.43%; IQR: 0.91% - 5.40%). Again, frequencies were similar for unexposed individuals (S1: 0.09%; IQR: 0.03% - 3.60%. S2: 1.48%; IQR: 0.46% - 2.26%). Representative examples of IFN-γ+ CD8+ T cells of a COVID-19 patient and an unexposed individual after short- and long-term stimulation is shown in Figure 4.

**Figure 4:**
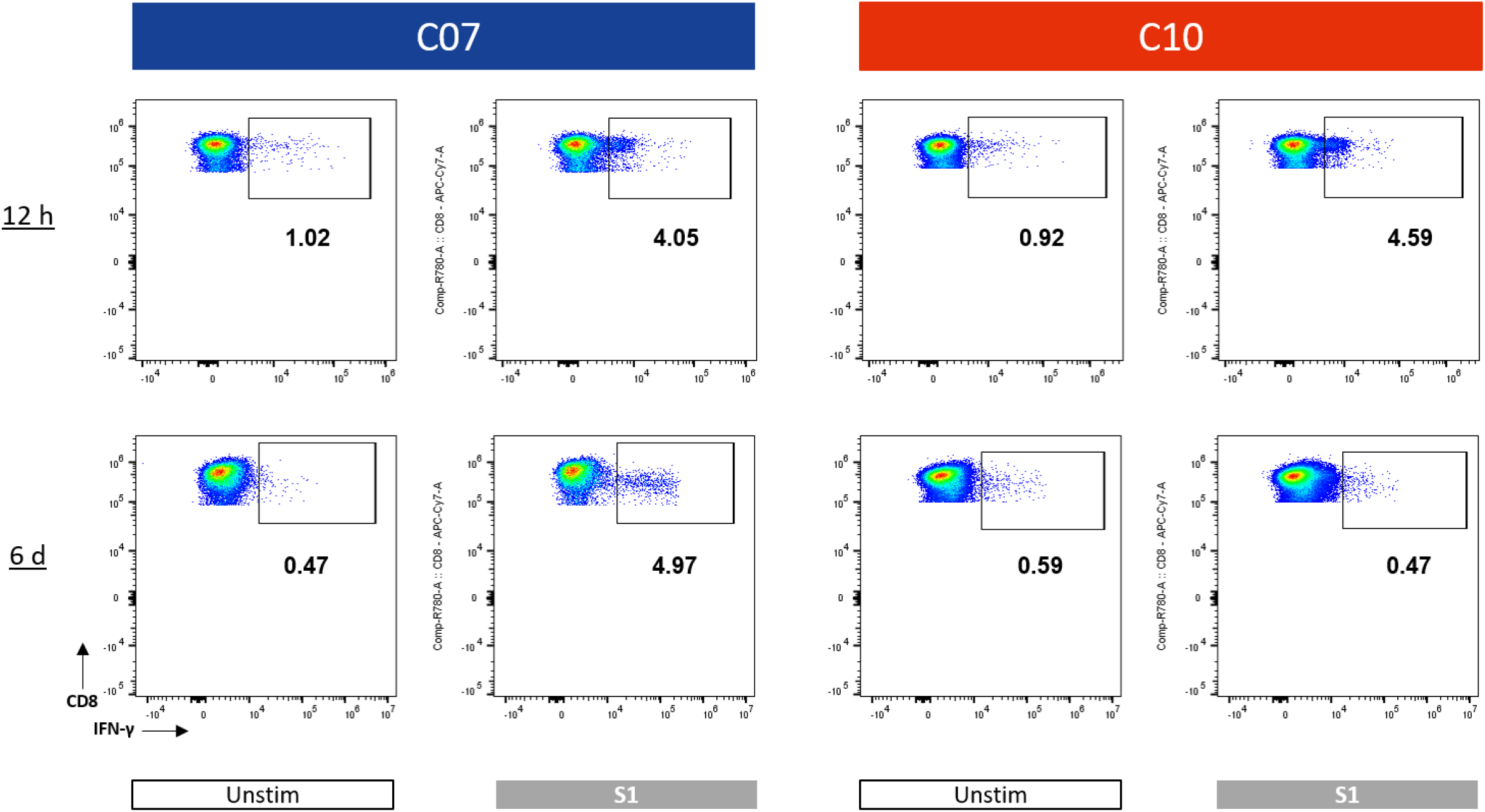
Representative examples of IFN-γ+ CD8+ T cells. IFN-γ+ CD8+ T cells of a COVID-19 patient (C07; see Table 1) and an unexposed individual (C10) after short- (upper row) and long-term (lower row) stimulation. Not restimulated (1^st^ and 3^rd^ column) and Spike1-restimulated cells (2^nd^ and 4^th^ column) are shown. Numbers indicate frequency of IFN-γ+ cells within all CD8+ T cells.

## Discussion

Our study confirms findings from previous reports showing that both CD4+ and CD8+ SARS-CoV-2-specific T cells can be detected in peripheral blood of recovered COVID-19 patients. In accordance with other groups, we observed that cellular immunity against SARS-CoV-2 is not only limited to the Spike protein but also targets the Nucleocapsid and Membrane protein. Strikingly, using our protocol, we did not detect SARS-CoV-2-specific T cells in unexposed donors.

Braun et al. detected Spike-reactive CD4+ T cells in 83% of COVID-19 patients, as well as in 34% of SARS-CoV-2 seronegative healthy donors. Cells were stimulated for 16 hours and antigen-specific T cells were detected using CD40L and 4-1BB expression (Braun et al. posted on medRxiv). Grifoni et al. detected CD4+ and CD8+ SARS-CoV-2-specific T cells in about 100% and 70% of COVID-19 convalescent patients, respectively. Targeted antigens included mainly Spike, Membrane and Nucleocapsid protein. SARS-CoV-2-reactive CD4+ T cells were also detected in 40% to 60% of unexposed individuals. CD4+ and CD8+ SARS- CoV-2-specific T cells were detected after stimulation for 24 hours and 9 hours, respectively (18). Oja et al. were able to detect CD4+ Spike-specific T cells in 21 of 21 COVID-19 patients with mild symptoms and 14 of 16 unexposed individuals. Cells were stimulated overnight and SARS-CoV-2-reactive T cells were identified by the upregulation of CD154 and 4-1BB (Oja et al. posted on bioRxiv). Gallais et al. detected SARS-CoV-2-specific T cells (targeting different antigens) in 9 of 9 COVID-19 patients and 6 of 8 relatives with neither a positive RT-qPCR results nor detectable antibodies against SARS-CoV-2. PBMCs were stimulated for about 20 hours and SARS-CoV-2-specific T cells were detected by an IFN-γ ELISPOT assay (Gallais et al. posted on medRxiv). Weiskopf et al. observed in 10 of 10 and 8 of 10 COVID-10 patients CD4+ and CD8+ T cells responses, respectively. SARS-CoV-2- reactive T cells were also detected in 2 out of 10 healthy controls not previously exposed to SARS-CoV-2. PBMCs were stimulated for about 20 hours and SARS-CoV-2-specific T cells were detected by an IFN-γ ELISPOT assay. PBMCs were stimulated for 20 hours and SARS-CoV-2-specific T cells were detected by the up-regulation of CD69 and CD137 (19).

The aforementioned studies have one aspect in common: PBMCs were stimulated for a rather short period of time. Viral epitopes are in principle much more immunogenic as compared to tumor neoantigens for example. Furthermore, especially the Spike protein is a very large protein (1273 amino acids), bearing a lot of potential class I and class II epitopes. The short incubation time might have led to a weak response of various naïve T cells or unspecific T cell responses. Interestingly, using the short-term assay, we observed especially Spike-specific T cells in unexposed individuals (8 of 9 individuals). Braun et al. observed that Spike-reactive T cells in COVID-19 expressed high levels of CD38 and HLA-DR. Two markers which are co-expressed on highly activated T cells. In contrast, Spike-reactive T cells of unexposed individuals did not express CD38 and HLA-DR or only at lower frequencies, probably reflecting their naïve origin (Braun et al. posted on medRxiv). Weiskopf et al. observed that SARS-CoV-2-specific CD4+ T cells of COVID-19 patients mainly showed a Central Memory phenotype, whereas CD8+ T cells mainly showed an Effector Memory or Terminally-differentiated Effector Memory phenotype. The phenotype of SARS-CoV-2- specific T cells of unexposed controls was not discussed (19).

In contrast, we were not able to detect SARS-CoV-2-reactive T cells in unexposed individuals. We might have been coincidentally selected individuals that have neither been exposed to SARS-CoV-2, nor to other common seasonal Coronaviruses. However, a short-term stimulation of PBMCs from the same unexposed donors led to significant immune responses probably indicating that the observed differences between our results and previously published data are not donor dependent. Peng et al. did also not see significant immune responses against SARS-CoV-2 antigens in healthy unexposed donors potentially reflecting the importance of experimental conditions (23).

Our data show that the Membrane protein might be an interesting target for studying in-depth humoral immune responses against SARS-CoV-2. A high frequency of recovered COVID-19 patients had Membrane-specific T cells in our small cohort. Intriguingly, the phenotype of these cells was almost exclusively restricted to the CD4 compartment. As T_Helper_ cells play an important role during B-cell priming, Membrane-specific antibodies might be present in these patients.

Deficiencies of our study include the small sample size and the focus on mild, non-hospitalized COVID-19 cases. With regard to this, we refrained from correlations between immunological and clinical meta-data. We have introduced a novel approach that can help to identify patients that were previously infected with SARS-CoV-2, so far with very high specificity. As compared to neutralization assays, our protocol does not require strict BSL3 safety conditions. Larger studies, correlating true-positive cellular and detailed humoral immune responses with clinical data might help to develop novel treatment strategies.

## Data Availability

All raw data can be obtained upon request

## Competing Interests

The authors have no competing interest.

## External funding

No external funding was received.

## Notes

### Competing Interest Statement

The authors have declared no competing interest.

### Author Declarations

All participants provided written informed consent. The study was approved by the Ethik-Kommission der Landesaerztekammer Baden-Wuerttemberg (F-2020-111).

